# Phase 1/2 Study to Describe the Safety and Immunogenicity of a COVID-19 RNA Vaccine Candidate (BNT162b1) in Adults 18 to 55 Years of Age: Interim Report

**DOI:** 10.1101/2020.06.30.20142570

**Authors:** Mark J. Mulligan, Kirsten E. Lyke, Nicholas Kitchin, Judith Absalon, Alejandra Gurtman, Stephen Lockhart, Kathleen Neuzil, Vanessa Raabe, Ruth Bailey, Kena A. Swanson, Ping Li, Kenneth Koury, Warren Kalina, David Cooper, Camila Fontes-Garfias, Pei-Yong Shi, Ӧzlem Türeci, Kristin R. Tompkins, Edward E. Walsh, Robert Frenck, Ann R. Falsey, Philip R. Dormitzer, William C. Gruber, Uğur Şahin, Kathrin U. Jansen

## Abstract

In March 2020, the WHO declared a pandemic of coronavirus disease 2019 (COVID-19), due to severe acute respiratory syndrome coronavirus 2 (SARS-CoV-2).^1^ With >8.8 million cases and >450,000 deaths reported globally, a vaccine is urgently needed. We report the available safety, tolerability, and immunogenicity data from an ongoing placebo-controlled, observer-blinded dose escalation study among healthy adults, 18-55 years of age, randomized to receive 2 doses, separated by 21 days, of 10 μg, 30 μg, or 100 μg of BNT162b1, a lipid nanoparticle-formulated, nucleoside-modified, mRNA vaccine that encodes trimerized SARS-CoV-2 spike glycoprotein RBD. Local reactions and systemic events were dose-dependent, generally mild to moderate, and transient. RBD-binding IgG concentrations and SARS-CoV-2 neutralizing titers in sera increased with dose level and after a second dose. Geometric mean neutralizing titers reached 1.8-to 2.8-fold that of a panel of COVID-19 convalescent human sera. These results support further evaluation of this mRNA vaccine candidate.

## Main

In December 2019, a pneumonia outbreak of unknown cause occurred in Wuhan, China. By January 2020, a novel coronavirus was identified as the etiologic agent. Within a month, the genetic sequence of the virus became available (MN908947.3). SARS-CoV-2 infections and the resulting disease, COVID-19, has spread globally. On 11 March 2020, the World Health Organization (WHO) declared the COVID-19 outbreak a pandemic. To date, the United States has reported the most cases globally.^2^ No vaccines are available to prevent SARS-CoV-2 infection or COVID-19 disease.

The RNA vaccine platform has enabled rapid vaccine development in response to this pandemic. RNA vaccines provide flexibility in the design and expression of vaccine antigens that can mimic antigen structure and expression during natural infection. RNA is required for protein synthesis, does not integrate into the genome, is transiently expressed, and is metabolized and eliminated by the body’s natural mechanisms and, therefore, is considered safe.^3,4,5,6^ RNA-based prophylactic infectious disease vaccines and RNA therapeutics have been shown to be safe and well-tolerated in clinical trials. In general, vaccination with RNA elicits a robust innate immune response. RNA directs expression of the vaccine antigen in host cells and has intrinsic adjuvant effects.^7^ A strength of the RNA vaccine manufacturing platform, irrespective of the encoded pathogen antigen, is the ability to rapidly produce large quantities of vaccine doses against a new pathogen. ^8,9^

Vaccine RNA can be modified by incorporating 1-methyl-pseudouridine which dampens innate immune sensing and increases mRNA translation *in vivo*.^10, 10^ The BNT162b1 vaccine candidate now being studied clinically incorporates such nucleoside modified RNA (modRNA) and encodes the receptor binding domain (RBD) of the SARS-CoV-2 spike protein, a key target of virus neutralizing antibodies. ^11^ The RBD antigen expressed by BNT162b1 is modified by the addition of a T4 fibritin-derived “foldon” trimerization domain to increase its immunogenicity^12^ by multivalent display.^13^ The vaccine RNA is formulated in lipid nanoparticles (LNPs) for more efficient delivery into cells after intramuscular injection.^14^ BNT162b1 is one of several RNA-based SARS-CoV-2 vaccine candidates being studied in parallel to select the candidate to advance to a safety and efficacy trial. Here, we present available data, through 14 days after a second dose in adults 18-55 years of age, from an ongoing Phase 1/2 vaccine study with BNT162b1, which is also being assessed in adults 65 to 85 years of age (ClinicalTrials.gov identifier: NCT04368728).

## Available Results

### Study Design and Demographics

Between 04 May 2020 and 19 June 2020, 76 subjects were screened, and 45 participants were randomized and vaccinated. Twelve participants per dose level (10 μg and 30 μg), were vaccinated with BNT162b1 on Days 1 and 21 and 12 participants received a 100 μg dose on Day 1. Nine participants received placebo (Figure 1). The study population consisted of healthy male and nonpregnant female participants with a mean age of 35.4 years (range 19 to 54 years); 51.1% were male and 48.9% were female. Most participants were white (82.2%) and non-Hispanic/non-Latino (93.3%) (Extended Data Table 1).

**Figure 1.**
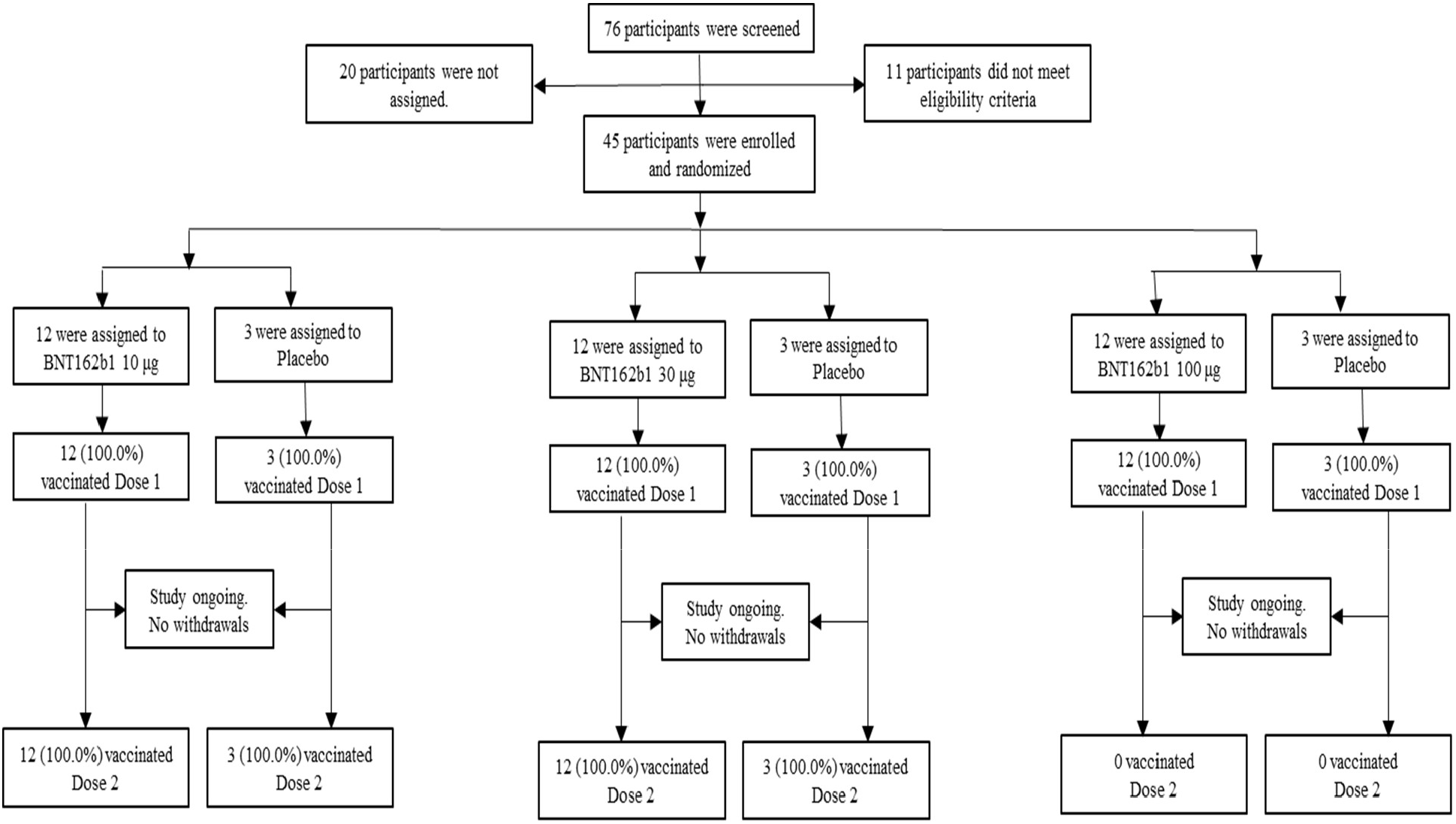
Disposition of participants. Participants not assigned (n-20) = participants who were screened but not randomized because enrollment had closed.

### Safety and Tolerability

In the 7 days following either Dose 1 or 2, pain at the injection site was the most frequent prompted local reaction, reported after Dose 1 by 58.3% (7/12) in the 10 μg, 100.0% (12/12 each) in the 30 μg and 100 μg BNT162b1 groups, and by 22.2% (2/9) of placebo recipients. After Dose 2, pain was reported by 83.3% and 100.0% of BNT162b1 recipients at the 10 μg and 30 μg dose levels, respectively, and by 16.7 % of placebo recipients. All local reactions were mild or moderate in severity except for one report of severe pain following Dose 1 of 100 μg BNT162b1 (Figure 2).

**Figure 2.**
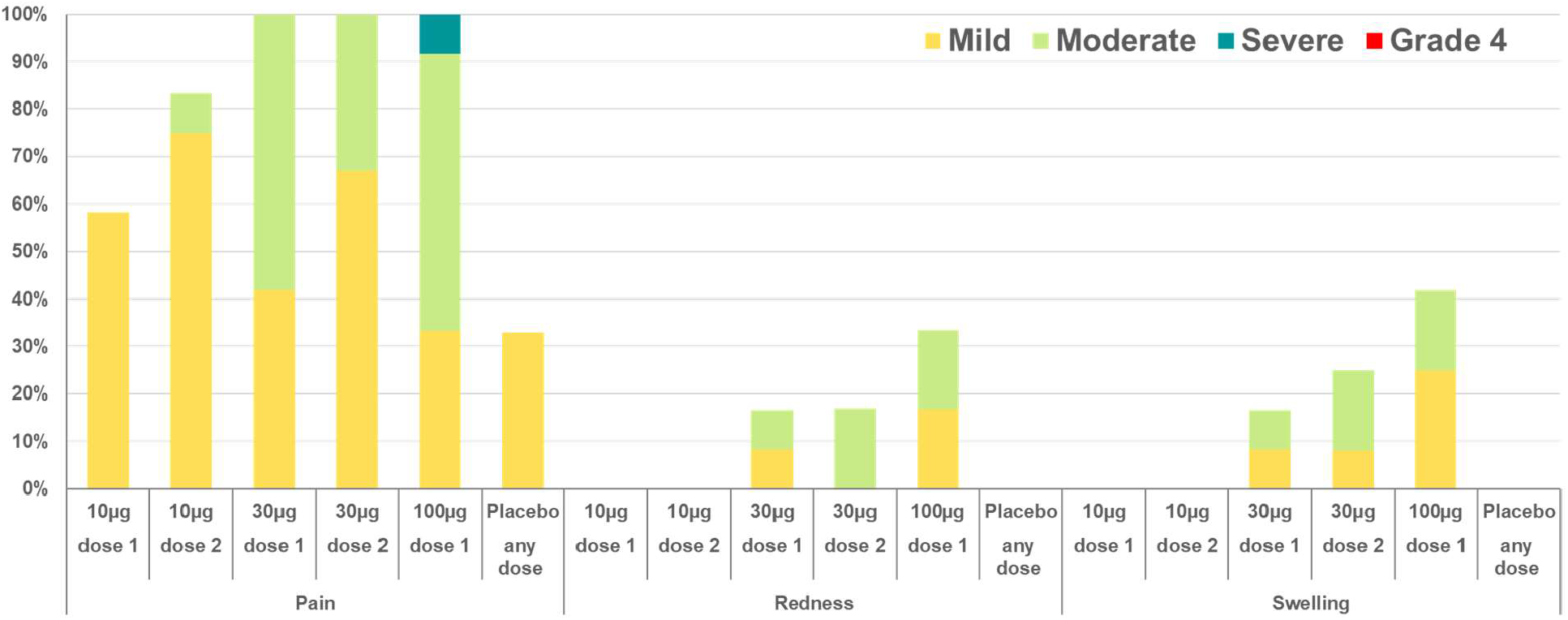
Local reactions reported within 7 days of vaccination, all dose levels. Solicited injection-site (local) reactions were: pain at injection site (mild: does not interfere with activity; moderate: interferes with activity; severe: prevents daily activity; Grade 4: emergency room visit or hospitalization) and redness and swelling (mild: 2.0 to 5.0 cm in diameter; moderate: >5.0 to 10.0 cm in diameter; severe: >10.0 cm in diameter; Grade 4: necrosis or exfoliative dermatitis for redness, and necrosis for swelling). Data were collected with the use of electronic diaries for 7 days after each vaccination.

The most common systemic events reported in the 7 days after each vaccination in both BNT162b1 and placebo recipients were mild to moderate fatigue and headache. Reports of fatigue and headache were more common in the BNT162b1 groups compared to placebo. Additionally, chills, muscle pain, and joint pain were reported among BNT162b1 recipients and not in placebo recipients. Systemic events increased with dose level and were reported in a greater number of subjects after the second dose (10 μg and 30 μg groups). Following Dose 1, fever (defined as ≥38.0 °C) was reported by 8.3% (1/12) of participants each in the 10 μg and 30 μg groups and in 50.0% (6/12) of BNT162b1 recipients in the 100 μg group. Based on the reactogenicity reported after the first dose of 100 μg, and the second dose of 30 μg, participants who received an initial 100 μg dose did not receive a second 100 μg dose. Following Dose 2, 8.3% (1/12) of participants in the 10 μg group and 75.0% (9/12) of participants in the 30 μg group reported fever ≥38.0 °C. Fevers generally resolved within 1 day of onset. No Grade 4 systemic events or fever were reported. (Figures 3a & 3b). Most local reactions and systemic events peaked by Day 2 after vaccination and resolved by Day 7.

**Figure 3.**
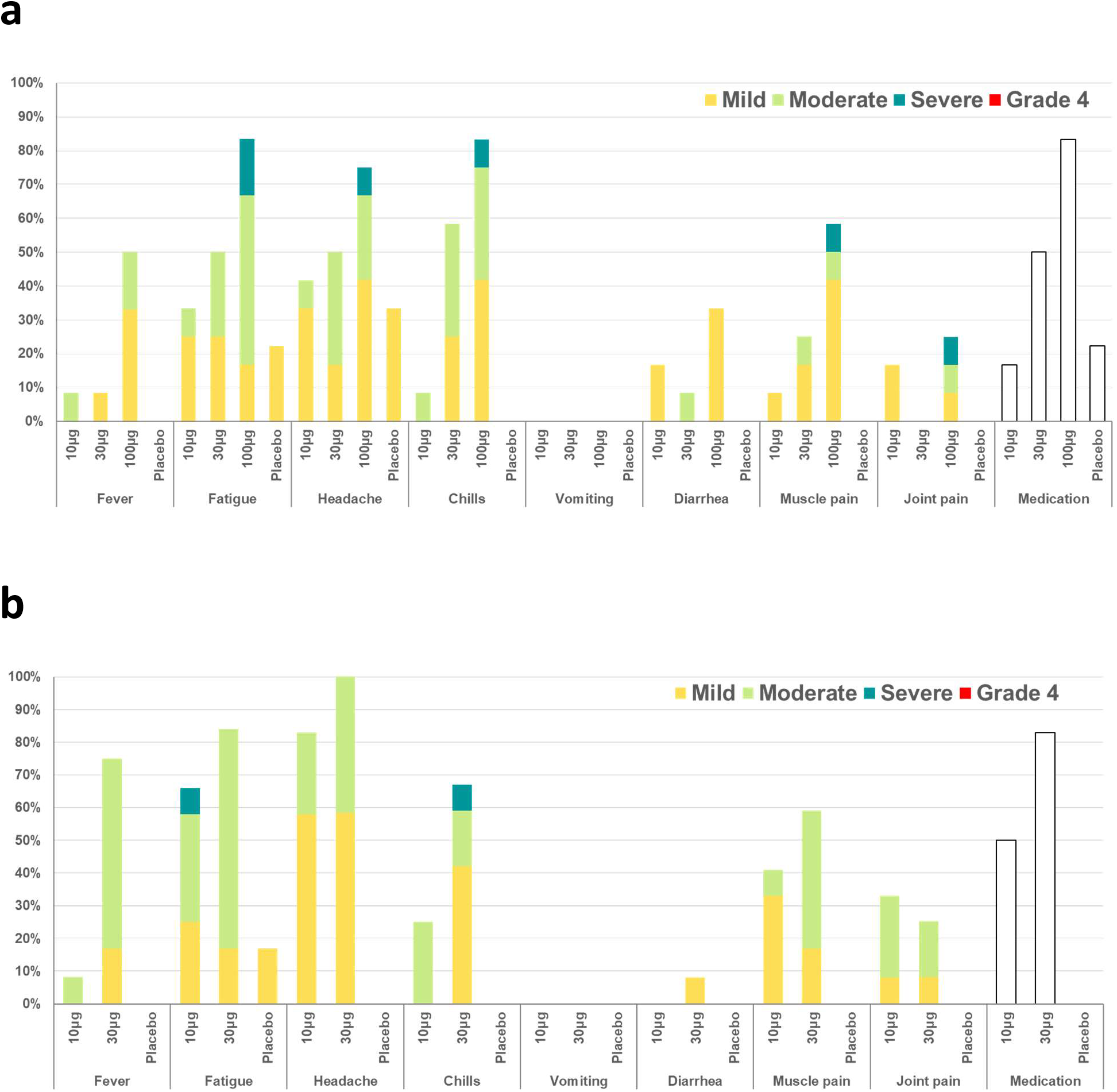
a. Systemic events and medication use reported within 7 days after vaccination 1, all dose levels and b. after vaccination 2, 10 μg and 30 μg dose levels. Solicited systemic events were: fatigue, headache, chills, new or worsened muscle pain, new or worsened joint pain (mild: does not interfere with activity; moderate: some interference with activity; severe: prevents daily activity), vomiting (mild: 1 to 2 times in 24 hours; moderate: >2 times in 24 hours; severe: requires intravenous hydration), diarrhea (mild: 2 to 3 loose stools in 24 hours; moderate: 4 to 5 loose stools in 24 hours; severe: 6 or more loose stools in 24 hours); Grade 4 for all events: emergency room visit or hospitalization; and fever (mild: 38.0°C to 38.4°C; moderate: 38.5°C to 38.9°C; severe: 39.0°C to 40.0°C; Grade 4: >40.0°C). Medication: proportion of participants reporting use of antipyretic or pain medication. Data were collected with the use of electronic diaries for 7 days after each vaccination.

Adverse events (Extended Data Table 1) were reported by 50.0% (6/12) of participants who received either 10 μg or 30 μg of BNT162b1, by 58.3% (7/12) of those who received 100 μg of BNT162b1, and by 11.1% (1/9) of placebo recipients. Two participants reported a severe adverse event: Grade 3 pyrexia 2 days after vaccination in the 30 μg group, and sleep disturbance 1 day after vaccination in the 100 μg group. Related AEs were reported by 25% (3/12 in the 10 μg groups) to 50% (6/12 each in 30 μg and 100 μg groups) of BNT162b1 recipients and by 11.1% (1/9) of placebo recipients. No serious adverse events were reported.

No Grade 1 or greater change in routine clinical laboratory values or laboratory abnormalities were observed for most subjects after either of the BNT162b1 vaccinations. Of those with laboratory changes, the most changes were decreases in lymphocyte count after Dose 1 in 8.3% (1/12), 45.5% (5/11), and 50.0% (6/12) of 10 μg, 30 μg, or 100 μg, recipients respectively, of BNT162b1. One participant each in the 10 μg group (8.3% [1/12]) and 30 μg group (9.1% [1/11]) dose levels and 4 participants at the 100 μg group (33.3% [4/12]) had Grade 3 decreases in lymphocytes. These post Dose 1 decreases in lymphocyte count, were transient and returned to normal 6-8 days after vaccination (Extended Data Figure 1). In addition, Grade 2 neutropenia was noted 6-8 days after the second dose of 10 μg or 30 μg BNT162b1, in 1 participant each. These two subjects continue to be followed in the study and no adverse events or clinical manifestation of neutropenia have been reported to date. None of the postvaccination abnormalities observed were associated with clinical findings.

### Immunogenicity

RBD-binding IgG concentrations and SARS-CoV-2 neutralizing titers were assessed at baseline and at 7 and 21 days after the first dose and 7 (Day 28) and 14 days (Day 35) after the second dose of BNT162b1 (Figure 4a). By 21 days after the first dose (for all three dose levels), geometric mean concentrations (GMCs) of RBD-binding IgG were 534-1,778 U/mL. In comparison, a panel of 38 SARS-CoV-2 infection/COVID-19 convalescent sera drawn at least 14 days after PCR-confirmed diagnosis from patients 18-83 years of age had an RBD-binding IgG GMC of 602 U/mL. By 7 days after the second dose (for the 10 μg and 30 μg dose levels) RBD-binding IgG GMCs had increased to 4,813-27,872 U/mL. RBD binding antibody concentrations among participants who received one dose of 100 μg BNT162b1 did not increase beyond 21 days after the first vaccination. In the participants who received the 10 μg and 30 μg doses of BNT162b1, highly elevated RBD-binding antibody concentrations persisted to the last time point evaluated (Day 35, 14 days after the second dose). These RBD-binding antibody concentrations were 5,880-16,166 U/mL compared to 602 U/mL in the human convalescent serum panel.

**Figure 4.**
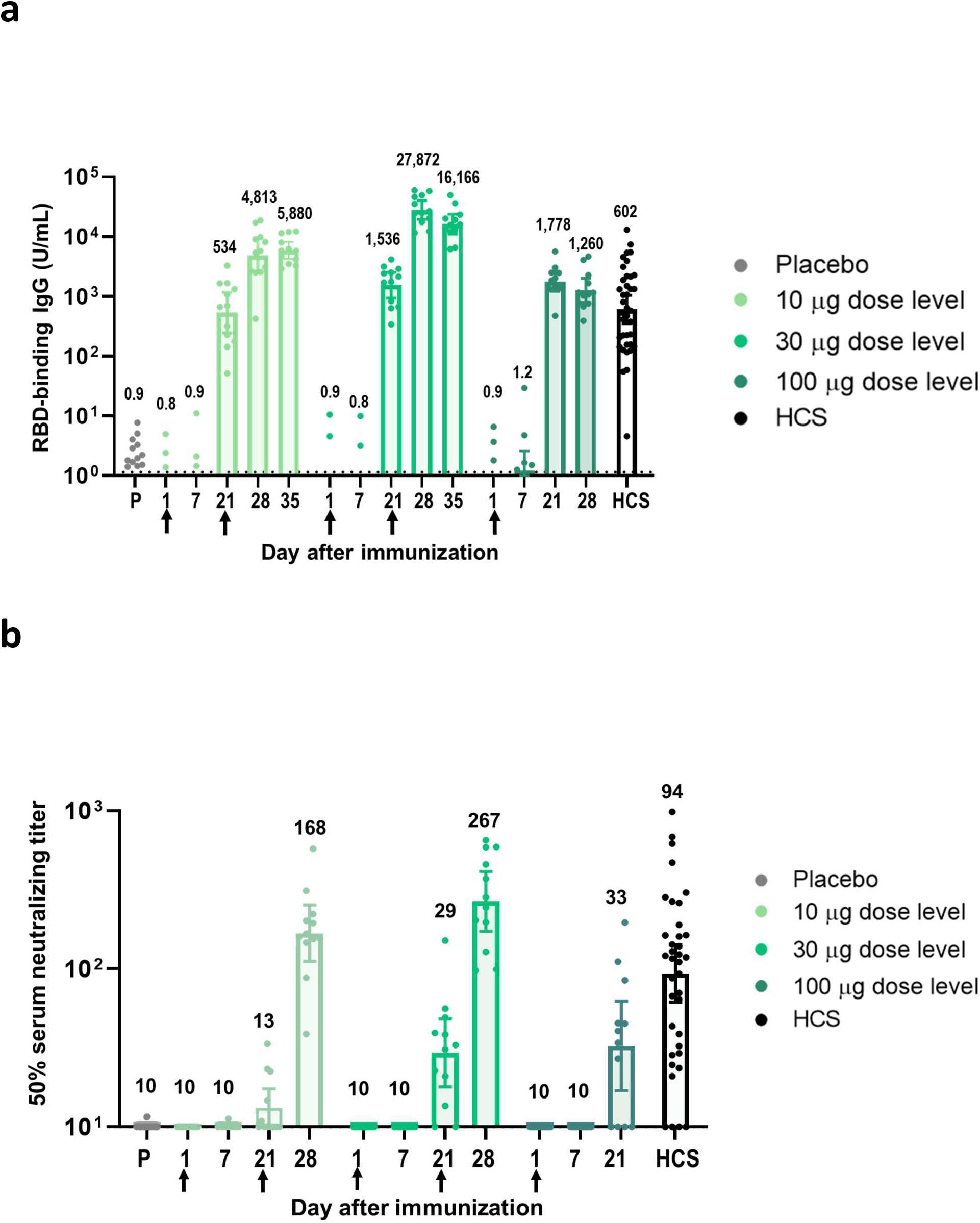
Immunogenicity of BNT162b1. Subjects in groups of 15 were immunized with the indicated dose levels of BNT162b1 (n=12) or with placebo (n=3) on days 1 (all dose levels and placebo) and 21 (10 μg and 30 μg dose levels and placebo). Reponses in placebo recipients are combined. The 28 day bleed is 7 days after the second immunization. Sera were obtained before immunization (Day 1) and 7, 21, and 28 days after the first immunization. Human COVID-19 convalescent sera (HCS, n=38) were obtained at least 14 days after PCR-confirmed diagnosis and at a time when the donors were asymptomatic. **a**. GMCs of recombinant RBD-binding IgG. Lower limit of quantitation (LLOQ) 1.15. **b**. 50% SARS-CoV-2 neutralizing GMTs. Each data point represents a serum sample, and each vertical bar represents a geometric mean with 95% confidence interval. The number above the bars are either the GMC or GMT for the group. Arrows indicate timing of vaccination (blood draws conducted prior to vaccination).

For all doses, modest increases in SARS-CoV-2 neutralizing geometric mean titers (GMTs) were observed 21 days after Dose 1 (Figure 4b). Substantially greater serum neutralizing GMTs were achieved 7 days after the second 10 μg or 30 μg dose, reaching 168-267, compared to 94 for the convalescent serum panel. The kinetics and durability of neutralizing titers are being monitored.

## Discussion

The RNA-based SARS-CoV-2 vaccine candidate BNT162b1 administered at 10 μg, 30 μg, and 100 μg to healthy adults 18-55 years of age exhibited a tolerability and safety profile consistent with those previously observed for mRNA-based vaccines.^5^ A clear dose-level response was observed after Doses 1 and 2 in adults 18-55 years of age. Based on the tolerability profile of the first dose at the 100 μg dose level and the second dose of 30 μg, participants randomized to the 100 μg group did not receive a second vaccination. Reactogenicity was generally higher after the second dose in the other two dosing levels, however symptoms were transient and resolved within a few days. Transient decreases in lymphocytes (Grades 1-3) were observed within a few days after vaccination, with lymphocyte counts returning to baseline within 6-8 days in all participants. These laboratory abnormalities were not associated with clinical findings. RNA vaccines are known to induce type I interferon which has been associated with transient migration of lymphocytes into tissues.^15, 16, 17,18^

Robust immunogenicity was observed after vaccination with BNT162b1. RBD-binding IgG concentrations were detected at 21 days after the first dose and substantially increased 7 days after the second dose given at Day 21. After the first dose, the RBD-binding IgG GMCs (10 μg dose recipients) were similar to those observed in a panel of 38 convalescent, human serology samples obtained at least 14 days after PCR-confirmed following SARS-CoV-2 infection/COVID-19 asymptomatic donors. Post-dose 1 GMCs were similar to those of the 30 μg and 100 μg groups but substantially higher than those in the convalescent serum panel. After Dose 2 with 10 μg or 30 μg BNT162b1, the RBD-binding IgG GMCs were ∼8.0-fold to ∼50-fold that of the convalescent serum panel GMC.

Neutralization titers were measurable after a single vaccination at Day 21 for all dose levels. At Day 28 (7 days after Dose 2), substantial SARS-CoV-2 neutralization titers were observed. The virus neutralizing GMTs after the 10 μg and 30 μg Dose 2 were, respectively, 1.8-fold and 2.8-fold the GMT of the convalescent serum panel. Assuming that neutralization titers induced by natural infection provide protection from COVID-19 disease, comparing vaccine-induced SARS-CoV-2 neutralization titers to those from sera of convalescent humans quantifies the magnitude of the vaccine-elicited response and the vaccine’s potential to provide protection. Since the 100 μg dose level cohort was not boosted, no corresponding data for immunogenicity after a second vaccination are available however there were no substantial differences in immunogenicity between the 30 μg and 100 μg dose levels after Dose 1. This observation suggests that a well-tolerated and immunogenic dose level may be between 10 μg and 30 μg for this vaccine candidate.

Our study had several limitations. While we used convalescent sera as a comparator, the kind of immunity (T cells versus B cells or both) and level of immunity needed to protect from COVID-19 are unknown. Further, this analysis of available data did not assess immune responses or safety beyond 2 weeks after the second dose of vaccine. Both are important to inform the public health use of this vaccine. Follow-up will continue for all participants and will include collection of serious adverse events for 6 months, and COVID-19 infection and multiple additional immunogenicity measurements through up to two years. While our population of healthy adults 55 years of age and younger is appropriate for a Phase 1/2 study, it does not accurately reflect the population at highest risk for COVID-19. Adults 65 years of age and over have already been enrolled in this study and results will be reported as they become available. Later phases of this study will prioritize enrollment of more diverse populations, including those with chronic underlying health conditions and from racial/ethnic groups adversely affected by COVID-19.^19^

These clinical findings for the BNT162b1 RNA-based vaccine candidate are encouraging and strongly support accelerated clinical development and at-risk manufacturing to maximize the opportunity for the rapid production of a SARS-CoV-2 vaccine to prevent COVID-19 disease.

## Role of the funding source

BioNTech is the Sponsor of the study. Pfizer was responsible for the design, data collection, data analysis, data interpretation, and writing of the report. The corresponding authors had full access to all the data in the study and had final responsibility for the decision to submit the data for publication. All study data were available to all authors.

## Data Sharing Statement

Upon request, and subject to review, Pfizer will provide the data that support the findings of this study. Subject to certain criteria, conditions and exceptions, Pfizer may also provide access to the related individual anonymized participant data. See https://www.pfizer.com/science/clinical-trials/trial-data-and-results for more information.

## Methods

### Study design

This study was conducted in healthy men and nonpregnant women 18 to 55 years of age to assess the safety, tolerability, and immunogenicity of ascending dose levels of various BNT162 mRNA vaccine candidates. In the part of the study reported here, assessment of three dose levels (10 μg, 30 μg, or 100 μg) of the BNT162b1 candidate was conducted at two sites in the United States. This study utilized a sentinel cohort design with progression and dose escalation taking place after review of data from the sentinel cohort at each dose level.

### Eligibility

Key exclusion criteria included individuals with known infection with human immunodeficiency virus, hepatitis C virus, or hepatitis B virus; immunocompromised individuals and those with a history of autoimmune disease; those with increased risk for severe COVID-19; previous clinical or microbiological diagnosis of COVID-19; receipt of medications intended to prevent COVID-19; previous vaccination with any coronavirus vaccine; a positive serological test for SARS-CoV-2 IgM and/or IgG at the screening visit; and a SARS-CoV-2 NAAT-positive nasal swab within 24 hours before study vaccination.

The final protocol and informed consent document were approved by institutional review boards for each of the participating investigational centers This study was conducted in compliance with all International Council for Harmonisation (ICH) Good Clinical Practice (GCP) guidelines and the ethical principles of the Declaration of Helsinki. A signed and dated informed consent form was required before any study-specific activity was performed.

### Endpoints

In this report, results from the following study primary endpoints are presented: the proportion of participants reporting prompted local reactions, systemic events, and use of antipyretic and/or pain medication within 7 days after vaccination, AEs and serious adverse events (SAEs) (available through up to ∼45 days after Dose 1), and the proportion of participants with clinical laboratory abnormalities 1 and 7 days after vaccination and grading shifts in laboratory assessments between baseline and 1 and 7 days after Dose 1 and between Dose 2 and 7 days after Dose 2. Secondary endpoints included: SARS-CoV-2 neutralizing geometric mean titers (GMTs); SARS-CoV-2 RBD-binding IgG geometric mean concentrations (GMCs) 7 and 21 days after Dose 1 and 7 and 14 days after Dose 2.

### Procedures

Study participants were randomly assigned to a vaccine group using an interactive web-based response technology system with each group comprising 15 participants (12 active vaccine recipients and 3 placebo recipients). Participants were to receive two 0.5-mL doses of either BNT162b1 or placebo, administered by intramuscular injection into the deltoid muscle.

BNT162b1 incorporates a Good Manufacturing Practice (GMP)-grade mRNA drug substance that encodes the trimerized SARS-CoV-2 spike glycoprotein RBD antigen. The mRNA is formulated with lipids as the mRNA-LNP drug product. The vaccine was supplied as a buffered-liquid solution for IM injection and was stored at −80 °C. The placebo was a sterile saline solution for injection (0.9% sodium chloride injection, in a 0.5-mL dose).

### Safety assessments

Safety assessments included a 4-hour observation after vaccination (for the first 5 participants vaccinated in each group), or a 30-minute observation (for the remainder of participants) for immediate AEs. The safety assessments also included self-reporting of prompted local reactions (redness, swelling, and pain at the injection site), systemic events (fever, fatigue, headache, chills, vomiting, diarrhea, muscle pain, and joint pain), and the use of antipyretic and/or pain medication in an electronic diary (e-diary) for 7 days after vaccination, and the reporting of unprompted AEs and SAEs after vaccination. Hematology and chemistry assessments were conducted at screening, 1 and 7 days after Dose 1, and 7 days after Dose 2.

There were protocol-specified safety stopping rules for all sentinel-cohort participants. Both an internal review committee (IRC) and an external data monitoring committee (EDMC) reviewed all safety data. No stopping rules were met prior to the publication of this report.

### Human convalescent serum panel

The 38 human SARS-CoV-2 infection/COVID-19 convalescent sera were drawn from subjects aged 18-83 years of age, at least 14 days after PCR-confirmed diagnosis, and at a time when subjects were asymptomatic. The serum donors predominantly had symptomatic infections (35/38), and one had been hospitalized. The sera were obtained from Sanguine Biosciences (Sherman Oaks, CA), the MT Group (Van Nuys, CA), and Pfizer Occupational Health and Wellness (Pearl River, NY).

### Immunogenicity assessments

50 mL of blood was collected for immunogenicity assessments before each study vaccination, at 7 and 21 days after Dose 1 and at 7 and 14 days after Dose 2. In the RBD-binding IgG assay, a recombinant SARS-CoV-2 RBD containing a C-terminal Avitag™ (Acro Biosystems) was bound to streptavidin-coated Luminex microspheres. Bound human anti-RBD antibodies were detected with a R-Phycoerythrin-conjugated goat anti-human polyclonal secondary antibody (Jackson Labs). Data were captured as median fluorescent intensities (MFIs) using a Luminex reader and converted to U/mL antibody concentrations using a reference standard curve with arbitrary assigned concentrations of 100 U/mL and accounting for the serum dilution factor. Assay results were reported in U/mL of IgG.

The SARS-CoV-2 neutralization assay used a previously described strain of SARS-CoV-2 (USA_WA1/2020) that had been rescued by reverse genetics and engineered by the insertion of an mNeonGreen (mNG) gene into open reading frame 7 of the viral genome.^20^ This reporter virus generates similar plaque morphologies and indistinguishable growth curves from wild-type virus. Viral master stocks used for the neutralization assay were grown in Vero E6 cells as previously described.^20^ Serial dilutions of heat inactivated sera were incubated with the reporter virus for 1 hour at 37°C before inoculating Vero CCL81 cell monolayers in 96 well plates to allow accurate quantification of infected cells. Total cell counts per well were enumerated by nuclear stain (Hoechst 33342) and fluorescent virally infected foci were detected 16-24 hours after inoculation with a Cytation 7 Cell Imaging Multi-Mode Reader (Biotek) with Gen5 Image Prime version 3.09. Titers were calculated in GraphPad Prism version 8.4.2 by generating a 4-parameter (4PL) logistical fit of the percent neutralization at each serial serum dilution. The 50% neutralization titer was reported as the interpolated reciprocal of the dilution yielding a 50% reduction in fluorescent viral foci.

### Statistical analysis

The sample size for the reported part of the study was not based on statistical hypothesis testing. The primary safety objective was evaluated by descriptive summary statistics for local reactions, systemic events, abnormal hematology and chemistry laboratory parameters, AEs, and SAEs after each vaccine dose for each vaccine group. The secondary immunogenicity objectives were descriptively summarized at the various time points. All participants with data available were included in the safety and immunogenicity analyses.

## Acknowledgements

The authors would like to thank Carol Monahan and Deb Gantt (Pfizer, Inc.) for writing and editorial support and Hua Ma, James Trammel, and Kiran Challagali (Pfizer Inc.) for statistical analysis support in the generation of this manuscript.

We would like to thank all the participants who volunteered for this study. We also acknowledge the following individuals for their contributions to this work:

NYU Langone Vaccine Center: Angelica Kottkamp, MD, Ramin Herati, MD, Rebecca Pellet Madan, MD, Mary Olson, DNP, ANP-BC, Marie Samanovic-Golden, PhD, Elisabeth Cohen, MD, Amber Cornelius, MS, Laura Frye, MPH, Heekoung Youn, RN, CCRC, MA, Baby Jane Fran, RN, Kanika Ballani, PharmD, MBA, Natalie Veling, RN, Juanita Erb, RN, BSN, MPA, Mahnoor Ali, BA, Lisa Zhao, MS, Stephanie Rettig, MPH, Hibah Khan, MPA, Harry Lambert, BA, Kelly Hu, BA, Jonathan Hyde, BS.

Center for Vaccine Development and Global Health, University of Maryland School of Medicine: Monica McArthur MD PhD, Justin Ortiz MD MS FACP FCCP, Rekha Rapaka MD, Linda Wadsworth RN, Ginny Cummings RN, Toni Robinson RN, Lisa Chrisley RN, Wanda Somrajit RN, Staci Eddington, Panagiota Komninou, Mardi Reymann, Kathy Strauss, Biraj Shrestha, Sudhaunshu Joshi, Robin Barnes RN, Roohali Sukhavasi, Alyson Kwon, Terry Sharp

University of Rochester and Rochester General Hospital: Emily Pierce, RN and Mary Criddle, RN

Cincinnati Children’s Hospital: Amy Cline RN, Susan Parker RN, Michelle Dickey APRN, Kristen Buschle APRN

Pfizer, Inc: Andrea Cawein, John L. Perez MD, MSc, Harpreet Seehra, Dina Tresnan DVM, PhD, Robert Maroko MD, Helen Smith, Sarah Tweedy, Amy Jones, Greg Adams, Rabia Malick, Emily Worobetz, Erica Weaver Liping Zhang, Carmel Devlin, Donna Boyce, Elisa Harkins Tull, Mark Boaz, Michael Cruz. Pfizer’s Clinical Assay Team.

BioNTech: Corinna Rosenbaum, Christian Miculka, Andreas Kuhn, Ferdia Bates, Paul Strecker, Alexandra Kemmer-Brück

## Author information

### Contributions

KUJ, PRD, WG, NK, SL, AG, RB, and US were involved in the design of the overall study and strategy. KN, MM, EW, RF, and AF provided feedback on the study design. WK, DC, KS, KT, CFG and PYS performed the immunological analyses. MJM, KN, EW, RF, AF, KL, and VR collected data as study investigators. PL and KK developed the statistical design and oversaw the data analysis. JA, KUJ, PRD, and WG drafted the initial version of the manuscript. All authors reviewed and edited the manuscript and approved the final version.

## Ethics declarations

### Competing interests

NK, JA, AG, SL, RB, KS, PL, KK, WK, DC, KT, PRD, WG, and KUJ are employees of Pfizer and may hold stock options. US and ÖT are stock owners, management board members, and employees at BioNTech SE (Mainz, Germany) and are inventors on patents and patent applications related to RNA technology. MJM, KL, KN, EW, AF, RF, and VR received compensation from Pfizer for their role as study investigators. CFG and PYS received compensation from Pfizer to perform the neutralization assay.

## Disclosures

These data are interim data from an ongoing study, database not locked. Data have not yet been source verified or subjected to standard quality check procedures that would occur at the time of database lock and may therefore be subject to change.

## Notes

### Clinical Trial

ClinicalTrials.gov identifier: NCT04368728

